# Temporal clinical and laboratory response to interleukin-6 receptor blockade with Tocilizumab in 89 hospitalized patients with COVID-19 pneumonia

**DOI:** 10.1101/2020.06.12.20122374

**Authors:** Daria S. Fomina, Mar’yana A. Lysenko, Irina P. Beloglazova, Zinaida Yu. Mutovina, Nataliya G. Poteshkina, Inna V. Samsonova, Tat’yana S. Kruglova, Anton A. Chernov, Alexander V. Karaulov, Michael M. Lederman

## Abstract

**Background:** Emerging evidence links morbidity and mortality of pandemic COVID-19 pneumonia to an inflammatory cytokine storm.

**Methods:** Eighty nine patients with COVID-19 pneumonia and heightened systemic inflammation (elevated serum C reactive protein and interleukin-6 levels) were treated with Tocilizumab (TCZ), a human monoclonal IgG1 antibody to the interleukin-6 receptor.

**Results:** Clinical and laboratory improvement was seen comparing baseline and 1-2 day post-infusion indices. Among 72 patients not receiving mechanical ventilation, NEWS2 scores fell from 5 to 2 (p < 0.001) C reactive protein levels fell from 95 to 14 mg/L (p < 0.001) and lymphocyte counts rose from 900 to 1000/uL (p = 0.036). Sixty three of 72 patients were discharged from hospital, one patient died, and 8 remained in hospital at time of writing. Among 17 patients receiving mechanical ventilation, despite a rapid decrease in CRP levels from 89 to 35 mg/L (p = 0.014) and early improvements in NEWS2 scores in 10 of 17, ten patients died and seven remain in hospital at time of writing. Overall, mortality was only seen in patients who had markedly elevated CRP levels (>30 mg/L) and low lymphocyte counts (< 1000/uL) before TCZ administration.

**Conclusions:** Inflammation and lymphocytopenia are linked to mortality in COVID-19. Inhibition of IL-6 activity by administration of Tocilizumab, an anti IL-6 receptor antibody is associated with rapid improvement in both CRP and lymphocyte counts and in clinical indices. Controlled clinical trials are needed to confirm the utility of IL-6 blockade in this setting. Additional interventions will be needed for patients requiring mechanical ventilation.

## INTRODUCTION

Currently, there are no approved effective treatments for patients with COVID-19. Clinical management of this infection is supportive with provision of supplementary oxygen and mechanical ventilation as warranted. Therefore, there is an urgent unmet need for effective treatment that will alter the morbid course of this infection and prevent mortality.

The COVID-19 pandemic is caused by infection with a new, readily transmissible RNA virus SARS-2. SARS-2 is a single-stranded coronavirus that enters cells after binding to the angiotensin converting enzyme type 2 (ACE-2) receptors that are highly expressed by nasal epithelium, type 2 alveolar cells and gut epithelial cells, but are also present in other organs^1,2,3^. While this virus can infect persons of all ages, morbidity and mortality are highest is among older persons and persons with underlying conditions such as obesity, cardiovascular disease, hypertension and diabetes^4,5^, who appear to be at greater risk for the development of pneumonia. Those severely ill with pneumonia develop an aggressive progression of disease that is characterized by elevated levels of inflammatory cytokines resembling in many ways a cytokine storm that has been described in other settings, such as for example, after administration chimeric antigen receptor (CAR) T cells acute lymphocyte leukemia^6^.

Here, we report our experience treating serious COVID-19 pneumonia with Tocilizumab (TCZ) a recombinant humanized monoclonal IgG1 antibody to the interleukin-6 receptor that binds both membrane and soluble receptors.

In this uncontrolled study of 89 persons with COVID-19 pneumonia who were admitted to a Moscow Territorial COVID Center, we found evidence of clinical improvement following TCZ administration as well as improvement in laboratory indices that are linked to COVD-19-induced cytokine storm.

## METHODS

### Patients and Intervention

Off-label administration of Tocilizumab was approved by the Local Ethics Committee of the 52^nd^ Moscow City Clinical Hospital for treatment of COVID-19 complicated by cytokine storm. We treated eighty-nine patients admitted to the Moscow City Clinical Hospital 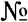 52 (a Territorial COVID Center) with a diagnosis of COVID-19 pneumonia. <u>SARS-CoV-2 infection</u> was confirmed by RT-PCR of nasopharyngeal swabs; pneumonia was diagnosed by CT scan. Tocilizumab 400 mg was administered intravenously based upon recent Russian Federal clinical recommendations for treatment of SARS-CoV-2 infection^7^.

To standardize initial and serial assessments of patients’ clinical status, the NEWS2 scale (the latest version of the National Early Warning Score) was used. NEWS2 scores were based on assessment of six physiological indices: respiratory rate; oxygen saturation; systolic blood pressure; heart rate; level of consciousness; temperature^8,9^.

### Laboratory and Radiology

Routine clinical laboratory assays were performed in the hospital clinical laboratories. Clinical and laboratory information was extracted from the patients’ medical records. C reactive protein (CRP) in plasma was measured by immunoturbidimetry (Beckman Coulter, Krefeld, Germany). Interleukin-6 (IL-6) levels in plasma were measured by electrochemiluminescence (Siemens Medical Solutions Diagnostics, Siemens Healthcare, Erlangen, Germany). CT Scans were read by experienced radiologists who scored results by severity using criteria as shown in Figure 1.

**Figure 1.**
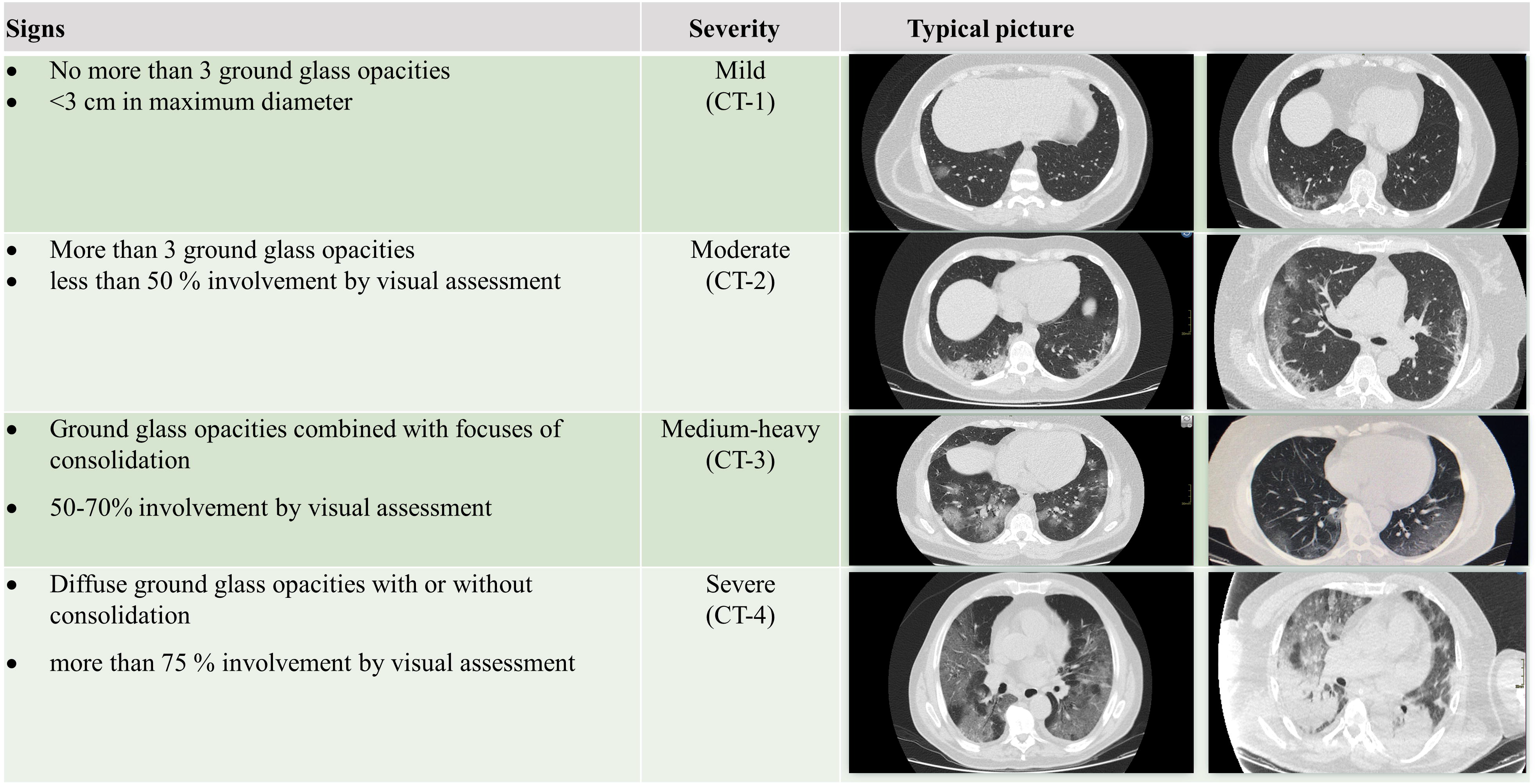
Clinical assessment of pneumonia severity based on computed tomography (CT) scores. Scoring method: Mild (CT-1) – no more than 3 ground glass opacities of < 3 cm in maximum diameter. Moderate (CT-2) – more than 3 ground glass opacities, less than 50 % involvement by visual assessment. Medium-heavy (CT-3) – ground glass opacities and pulmonary consolidation, 50–70% involvement by visual assessment. Severe (CT-4) – diffuse ground glass opacities with or without consolidation, more than 75 % involvement by visual assessment.

### Statistical analysis

Data are presented as medians and interquartile ranges (IQR).

Statistical data processing was carried out using the application program “IBM SPSS STATISTICS V-22” software. For comparison of quantitative indicators, the Mann-Whitney U test was used, and for qualitative characteristics, the Fisher χ2 test was used. Differences were considered significant if p < 0.05.

## RESULTS

Clinical characteristics of the 89 patients before administration of TCZ are shown in Table 1. Seventeen of these patients (19%) were receiving mechanical ventilation (MV) while 72 (81%) were treated with supplemental oxygen without mechanical ventilation (No MV). Ten of these 72 patients were receiving high flow oxygen.

**Table 1.**
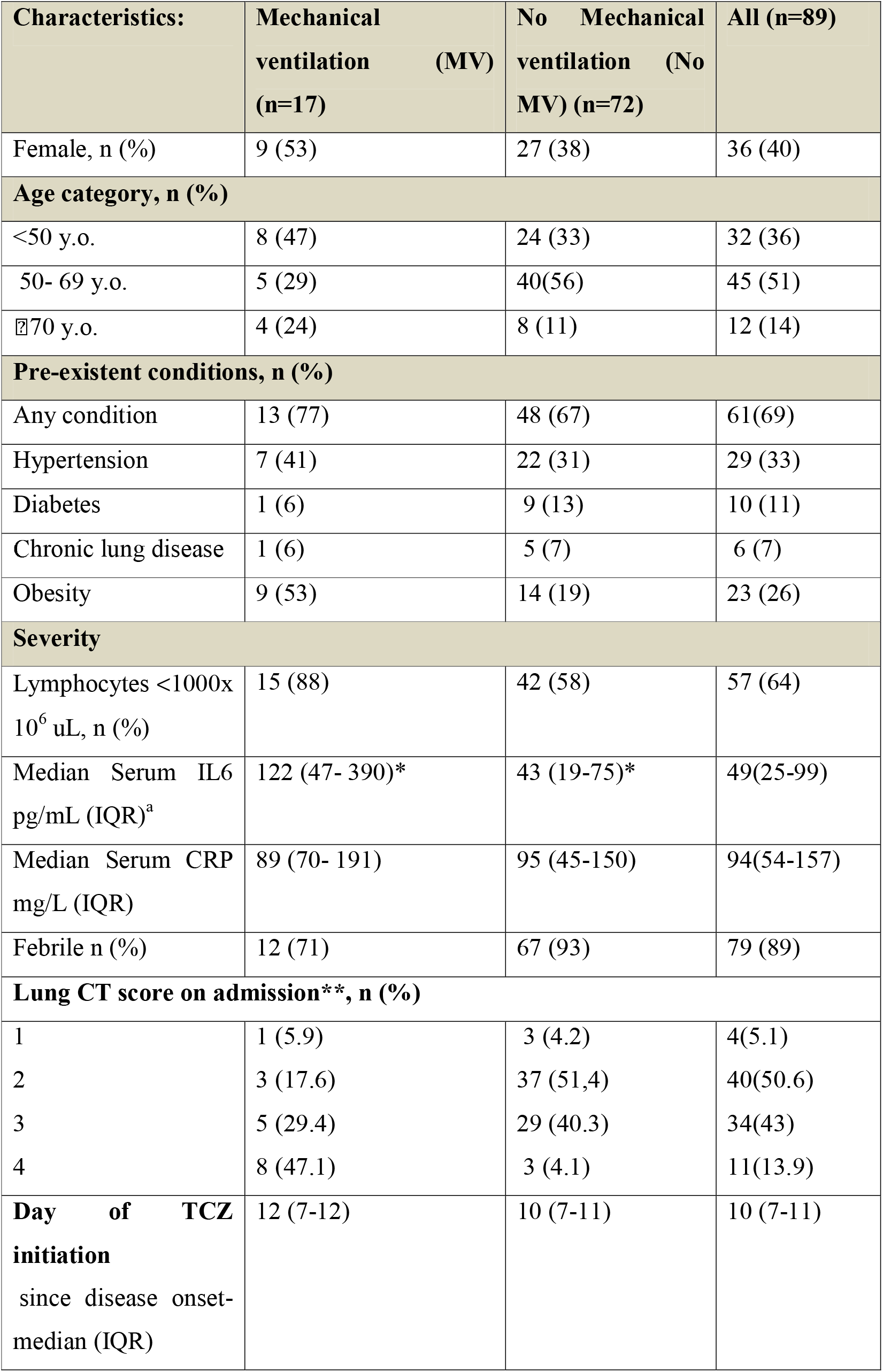
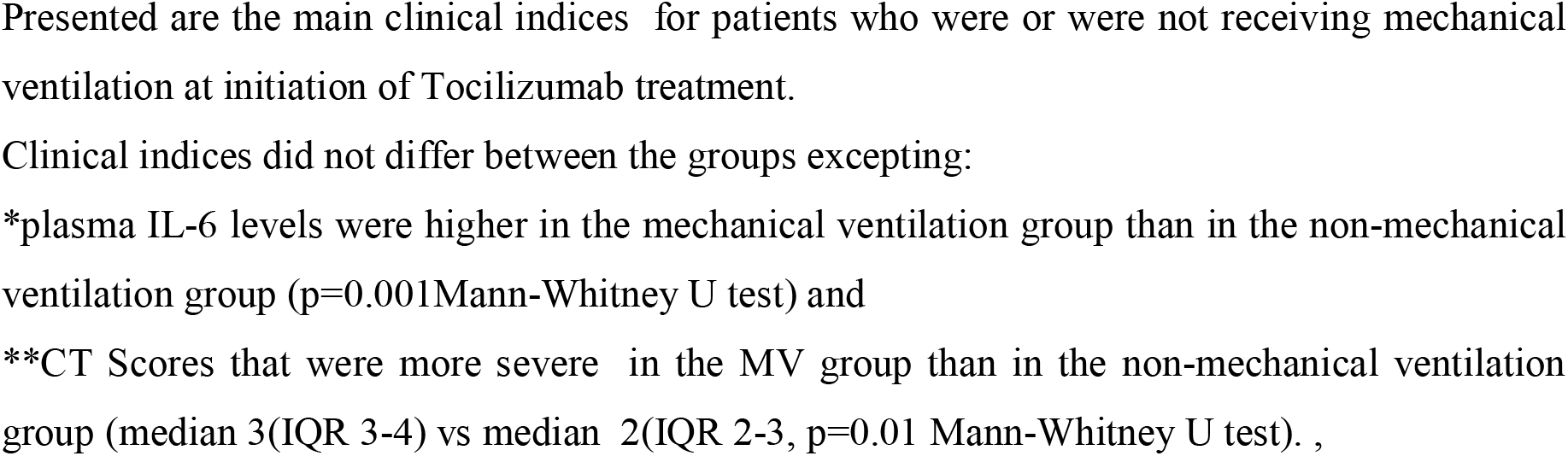
Clinical characteristics of patients before administration of Tocilizumab.

| **Characteristics:** | **Mechanical ventilation (MV) (n=17)** | **No Mechanical ventilation (No MV) (n=72)** | **All (n=89)** |
| --- | --- | --- | --- |
| Female, n (%) | 9 (53) | 27 (38) | 36 (40) |
| **Age category, n (%)** |  |  |  |
| <50 y.o. | 8 (47) | 24 (33) | 32 (36) |
| 50- 69 y.o. | 5 (29) | 40(56) | 45 (51) |
| ≥70 y.o. | 4 (24) | 8 (11) | 12 (14) |
| **Pre-existent conditions, n (%)** |  |  |  |
| Any condition | 13 (77) | 48 (67) | 61(69) |
| Hypertension | 7 (41) | 22 (31) | 29 (33) |
| Diabetes | 1 (6) | 9 (13) | 10 (11) |
| Chronic lung disease | 1 (6) | 5 (7) | 6 (7) |
| Obesity | 9 (53) | 14 (19) | 23 (26) |
| **Severity** |  |  |  |
| Lymphocytes <1000x 10<sup>6</sup> uL, n (%) | 15 (88) | 42 (58) | 57 (64) |
| Median Serum IL6 pg/mL (IQR)<sup>a</sup> | 122 (47- 390)* | 43 (19-75)* | 49(25-99) |
| Median Serum CRP mg/L (IQR) | 89 (70- 191) | 95 (45-150) | 94(54-157) |
| Febrile n (%) | 12 (71) | 67 (93) | 79 (89) |
| **Lung CT score on admission\*\*, n (%)** |  |  |  |
| 1 | 1 (5.9) | 3 (4.2) | 4(5.1) |
| 2 | 3 (17.6) | 37 (51,4) | 40(50.6) |
| 3 | 5 (29.4) | 29 (40.3) | 34(43) |
| 4 | 8 (47.1) | 3 (4.1) | 11(13.9) |
| **Day of TCZ initiation**<br>since disease onset-<br>median (IQR) | 12 (7-12) | 10 (7-11) | 10 (7-11) |

Presented are the main clinical indices for patients who were or were not receiving mechanical ventilation at initiation of Tocilizumab treatment.
Clinical indices did not differ between the groups excepting:
\*plasma IL-6 levels were higher in the mechanical ventilation group than in the non-mechanical ventilation group ( $p=0.001$ Mann-Whitney U test) and
\*\*CT Scores that were more severe in the MV group than in the non-mechanical ventilation group (median 3(IQR 3-4) vs median 2(IQR 2-3, $p=0.01$ Mann-Whitney U test). ,

Slightly over half of the patients in the MV group were 50 years old or older while 2/3 of patients who were in the No MV group were 50 years old. or older. Half of the MV patients were women while only approximately 1/3 (27/92) No MV patients were women. None of these demographics were significantly different between the MV and No MV groups. Pre-existent comorbidities were present in 77% patients (3/17) of the MV group and 67% (48/72) of the No MV group (no statistical difference, NSD). Hypertension was present in 41% of the MV group and in 31% of the No MV group (NSD) and obesity in 53% of the MV group and 19% of the No MV group (p = 0.04). Underlying diabetes was seen in only 1/17 MV patients and in 9/72 No MV patients and chronic lung disease was in 1/17 MV patients and 5/72 No MV patients (NSD).

Circulating lymphocyte numbers were low (< 1000/uL) in 88% of MV patients (15/17) and in 58% of the No MV patients (42/72). Serum IL-6 levels were elevated and were higher in the MV group with median levels of 122 pg/mL (IQR 47–390) than in the No MV group (48 pg/mL (IQR 19–75, p = 0.001)). Serum levels of C Reactive Protein (CRP) an acute phase reactant that is driven by IL-6 were elevated in both groups with medians of 89 mg/L (IQR 70–191) in the MV group and 95 mg/L (IQR 45–150) in the No MV group (NSD). CT scores were more severe in the MV group with median scores of 3 versus median scores of 2 in the No MV group (p = 0.01). Nearly half of patients (8/17) in the MV group had scores of 4 while in the No MV group, only 3 of 72 (4%) had scores of 4. Among MV patients, TCZ administration was begun on median day 12 of illness (IQR 7–12 days) and on median 10^th^ day of illness (IQR 7–11 days) among No MV patients.

Among the 89 patients who were treated with TCZ, 74 had been treated for a median of 9 days with hydroxychloroquine+ azithromycin + lopinavir/ritonavir before TCZ treatment, 4 had been treated for a median of 9 days with hydroxychloroquine+azithromycin before TCZ treatment and 11 had been treated for a median of 9 days with lopinavir/ritonavir before TCZ treatment.

Figure 2 shows the changes in clinical status as determined by NEWS 2 scores, from baseline at the initiation of Tocilizumab to 1–2 days after TCZ administration. In the No MV group, median NEWS2 scores fell from 5 to 2 (p < 0.001, Mann-Whitney U test) with 65/72 patients experiencing a decrease reflecting clinical improvement, 6/72 remaining unchanged, and 1/72 increasing score reflecting clinical deterioration. None required mechanical ventilation. In the MV group, median NEWS2 scores remained at 8, with 10/17 experiencing a decrease reflecting improvement, 4/17 remaining unchanged, and 3/17 experiencing an increase in scores reflecting deterioration. Laboratory results at baseline before TCZ administration and 1–2 days after treatment are shown in Table 2. CRP levels fell in the No MV group from a median of 95 mg/L pre-treatment to 13.5 mg/L 1–2 days after treatment (p < 0.001). CRP levels also fell in the MV group from 89 mg/L before TCZ administration to 35 mg/L 1–2 days after TCZ treatment (p = 0.014). Absolute lymphocyte counts rose from a median of 900/uL to 1000/uL (p = 0.036) in the No MV group, but there was not a significant change in the MV group (from 700 to 800 cells/uL). In both groups, LDH, AST, ALT and fibrinogen levels did not change significantly although sampling was incomplete. The absolute leukocyte count increased significantly in the MV group from 5,800/uL to 9,100/uL (p = 0.02), while in the No MV group there was a non-significant decrease from 5,500/uL to 4,000/uL. The absolute neutrophil count increased after TCZ treatment in the MV group from 3,900 /uL to 7,400/uL (p = 0.001) while a decrease from 4,200/uL to 2,800/uL in the No MV group was not significant.

**Figure 2.**
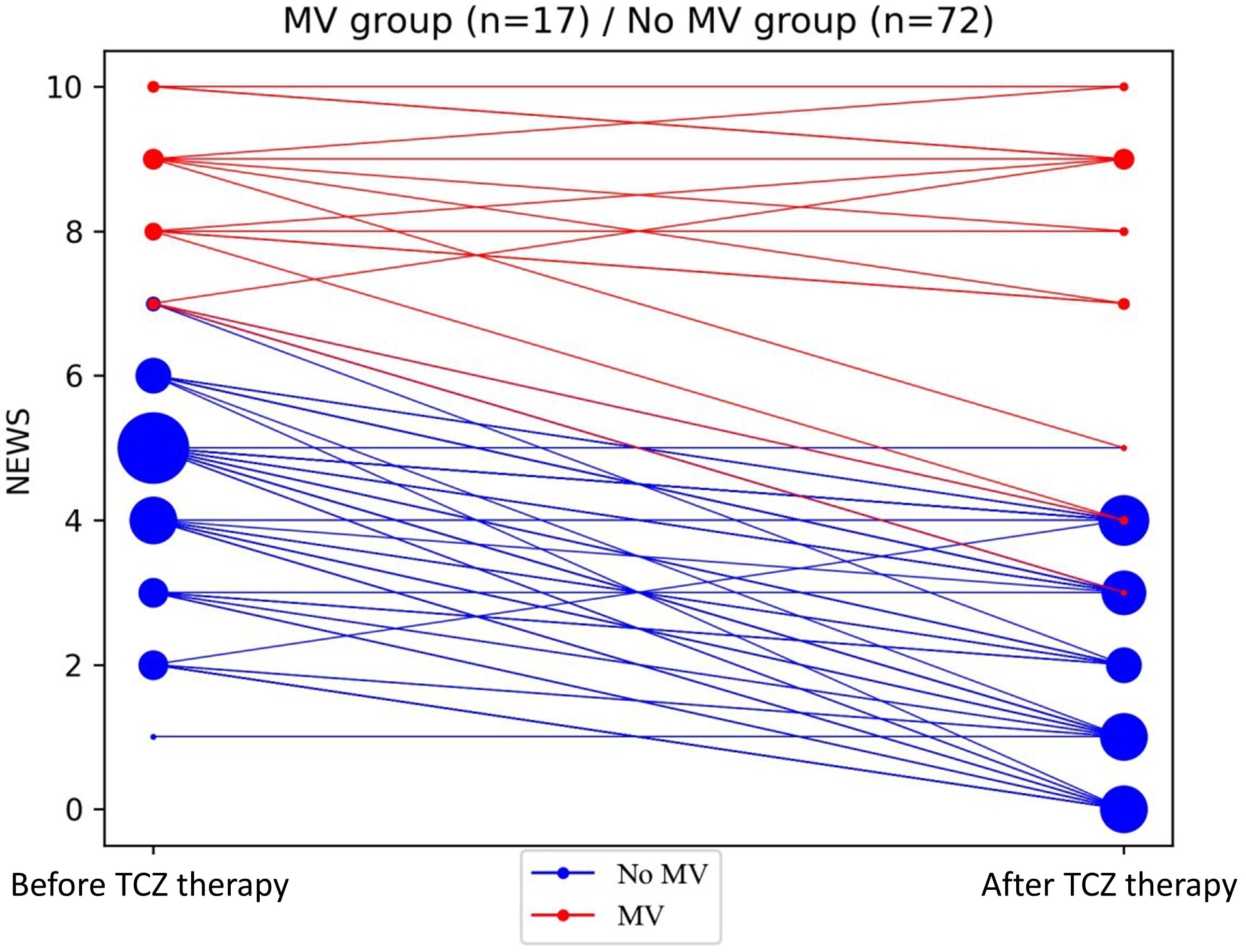
Changes in NEWS2 severity score after TCZ therapy. NEWS2 severity score is shown before (day of TCZ initiation) and 24–48 h after launching TCZ therapy for 17 patients who were receiving mechanical ventilation (MV) (red) or who were not receiving mechanical ventilation (blue)prior to therapy. The size of the circle reflects the number of patients with a given severity score.

**Table 2.**
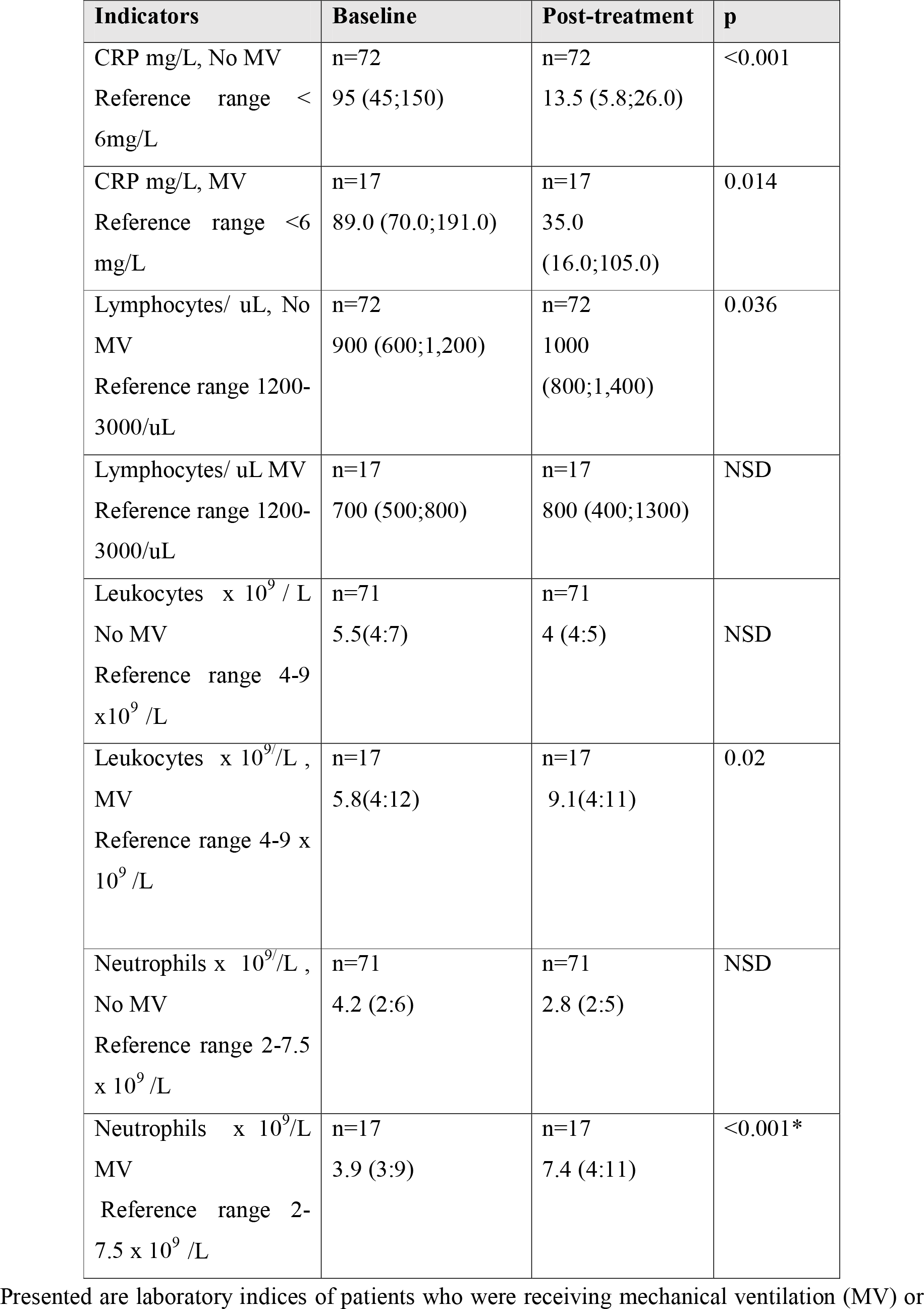
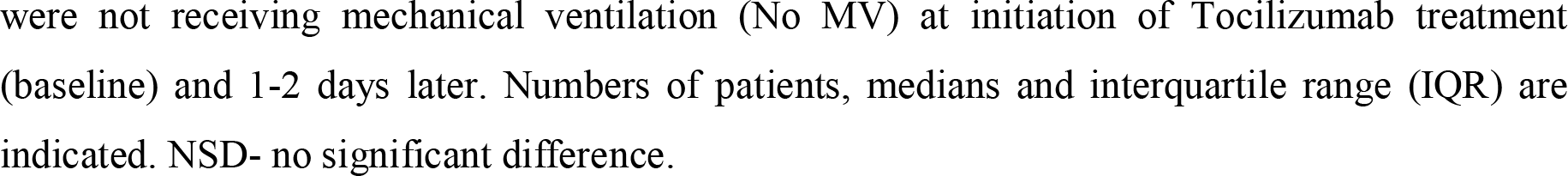
Early changes in laboratory indices after TCZ.

| Indicators | Baseline | Post-treatment | p |
| --- | --- | --- | --- |
| CRP mg/L, No MV<br>Reference range < 6mg/L | n=72<br>95 (45;150) | n=72<br>13.5 (5.8;26.0) | <0.001 |
| CRP mg/L, MV<br>Reference range <6 mg/L | n=17<br>89.0 (70.0;191.0) | n=17<br>35.0<br>(16.0;105.0) | 0.014 |
| Lymphocytes/ uL, No MV<br>Reference range 1200-3000/uL | n=72<br>900 (600;1,200) | n=72<br>1000<br>(800;1,400) | 0.036 |
| Lymphocytes/ uL MV<br>Reference range 1200-3000/uL | n=17<br>700 (500;800) | n=17<br>800 (400;1300) | NSD |
| Leukocytes $\times 10^9 / L$<br>No MV<br>Reference range 4-9 $\times 10^9 / L$ | n=71<br>5.5(4:7) | n=71<br>4 (4:5) | NSD |
| Leukocytes $\times 10^9 / L$ ,<br>MV<br>Reference range 4-9 $\times 10^9 / L$ | n=17<br>5.8(4:12) | n=17<br>9.1(4:11) | 0.02 |
| Neutrophils $\times 10^9 / L$ ,<br>No MV<br>Reference range 2-7.5 $\times 10^9 / L$ | n=71<br>4.2 (2:6) | n=71<br>2.8 (2:5) | NSD |
| Neutrophils $\times 10^9 / L$<br>MV<br>Reference range 2-7.5 $\times 10^9 / L$ | n=17<br>3.9 (3:9) | n=17<br>7.4 (4:11) | <0.001* |

Presented are laboratory indices of patients who were receiving mechanical ventilation (MV) or
were not receiving mechanical ventilation (No MV) at initiation of Tocilizumab treatment (baseline) and 1-2 days later. Numbers of patients, medians and interquartile range (IQR) are indicated. NSD- no significant difference.

### Clinical Outcomes

At the time of data set freezing, between days 6 and 26 after TCZ administration, 63 patients had been discharged from the hospital, 11 died, while 15 patients remained in the hospital (Figure 3). In the No MV group, one patient died (1%), 63 patients were discharged from the hospital (88%), and 8 patients (11%) remained in the hospital at a median of 11days (range 6–23) after TCZ administration. In the MV group, 10 patients died (59%), 7 patients remained in hospital at a median 13 days (range 9–24) after TCZ administration, and no patients were discharged from the hospital. (Figure 3).

**Figure 3.**
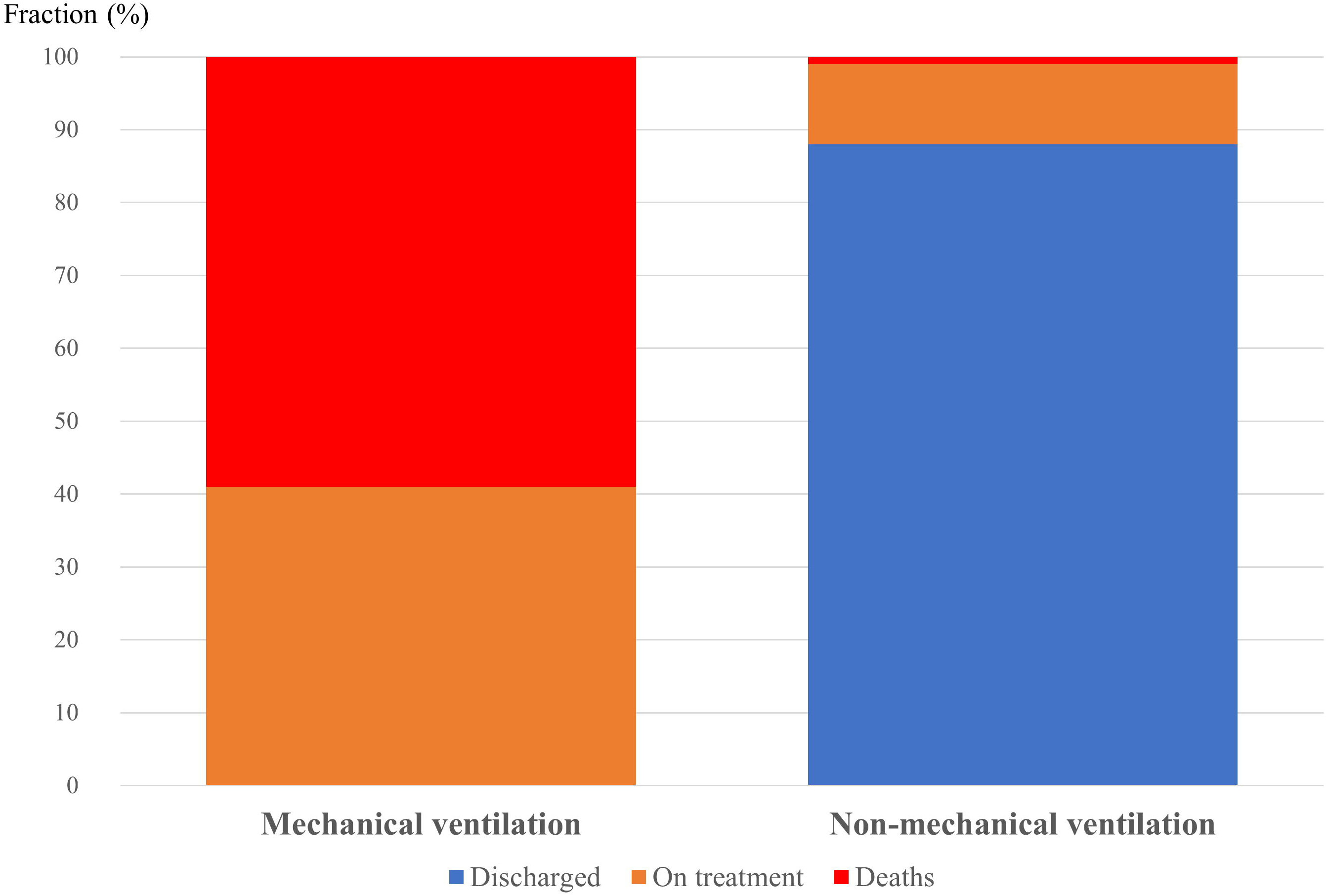
Clinical outcomes according to use of mechanical ventilation before TCZ administration. Outcomes are shown for the 17 patients who were on mechanical ventilation and 62 patients who were not on mechanical ventilation before TCZ therapy. Proportion of deaths are in red, proportions of patients still in hospital are in orange and proportions discharged from hospital in blue. Mortality was greater in patients receiving mechanical ventilation p < 0.001 Fisher χ2 test).

Combined data on clinical and laboratory indices among patients who died (n = 11) and those who survived and were discharged (n = 63) in the MV and No MV groups, are presented in Table 3. Patients who remained on treatment in the hospital were excluded from this analysis. Median baseline NEWS2 scores were higher in those who died than those who survived (9 vs 4; p< 0.001), as were median NEWS2 scores 1–2 days after treatment with TCZ (9 vs 2; p< 0.001). Median baseline CRP levels tended to be higher in those who died than in those who survived (168 vs 94 mg/L), however these differences did not reach statistical significance (p = 0.059). After TCZ treatment, median CRP levels fell (p< 0.001) in both groups but remained higher in those who died than in those who survived (51 vs 13 mg/L, p = 0.001). Baseline lymphocyte counts were lower in lethal cases than in survivors (600 vs 900/uL, p = 0.003) and remained lower in fatal cases than in survivors after treatment with TCZ (800 vs 1100/uL, p = 0.05). Baseline and after-treatment levels of LDH were not different between these groups (data not shown).

**Table 3.** Comparison of clinical and laboratory indices in patients who recovered or died.

| **Index** | **Death** | **Recovery**<br>**(discharge)** | **p** |
| --- | --- | --- | --- |
| NEWS2 baseline | n=11;<br>9 (8: 10) | n=63;<br>4(3:5) | <0.001* |
| NEWS2, after TCZ | n=11<br>9 (8: 9) | n=63<br>2(1: 3) | <0.001* |
| CRP mg/L baseline | n=11<br>168 (77: 205) | n=63<br>94 (33:148) | NSD |
| CRP mg/L, after TCZ | n=11<br>51(23: 153) | n=63<br>13 (5: 23) | 0.001 |
| Lymphocytes /uL baseline | n=11<br>600 (500: 700) | n=63<br>900 (700: 1200) | 0.003 |
| Lymphocytes /uL, after TCZ | n=11<br>800 (350: 1100) | n=63<br>1100 (850: 1400) | 0.05 |

Presented are clinical and laboratory indices at baseline (just before TCZ administration) and one-to-two days after TCZ treatment in patients who died and patients who survived to discharge from hospital. Number of patients (n) and interquartile ranges (IQR) are presented. NSD -no statistical differences Data for total of 74 patients. Another 15 patients remain in hospital, at time of writing.

We found that absolute lymphopenia and elevated CRP levels immediately before treatment with TCZ could reliably distinguish survivors from those patients who died. Shown in Figure 4 are the survival curves for patients with known outcomes divided into four groups according to absolute lymphocyte counts above or lower than 1000^/^/uL at baseline and CRP levels lower or greater than 30 mg/L at baseline. There were 41 patients in the highest CRP, low lymphocyte count group, 22 patients in the highest CRP normal lymphocyte count group, 5 patients in the lower CRP low lymphocyte group, and 6 patients in the lower CRP normal lymphocyte count group. Mortality was only seen in patients who had both an absolute lymphocyte count < 1000/uL and a CRP > 30 mg/L. Lymphopenia alone or elevated CRP levels without lymphopenia were not seen in fatal cases.

**Figure 4.**
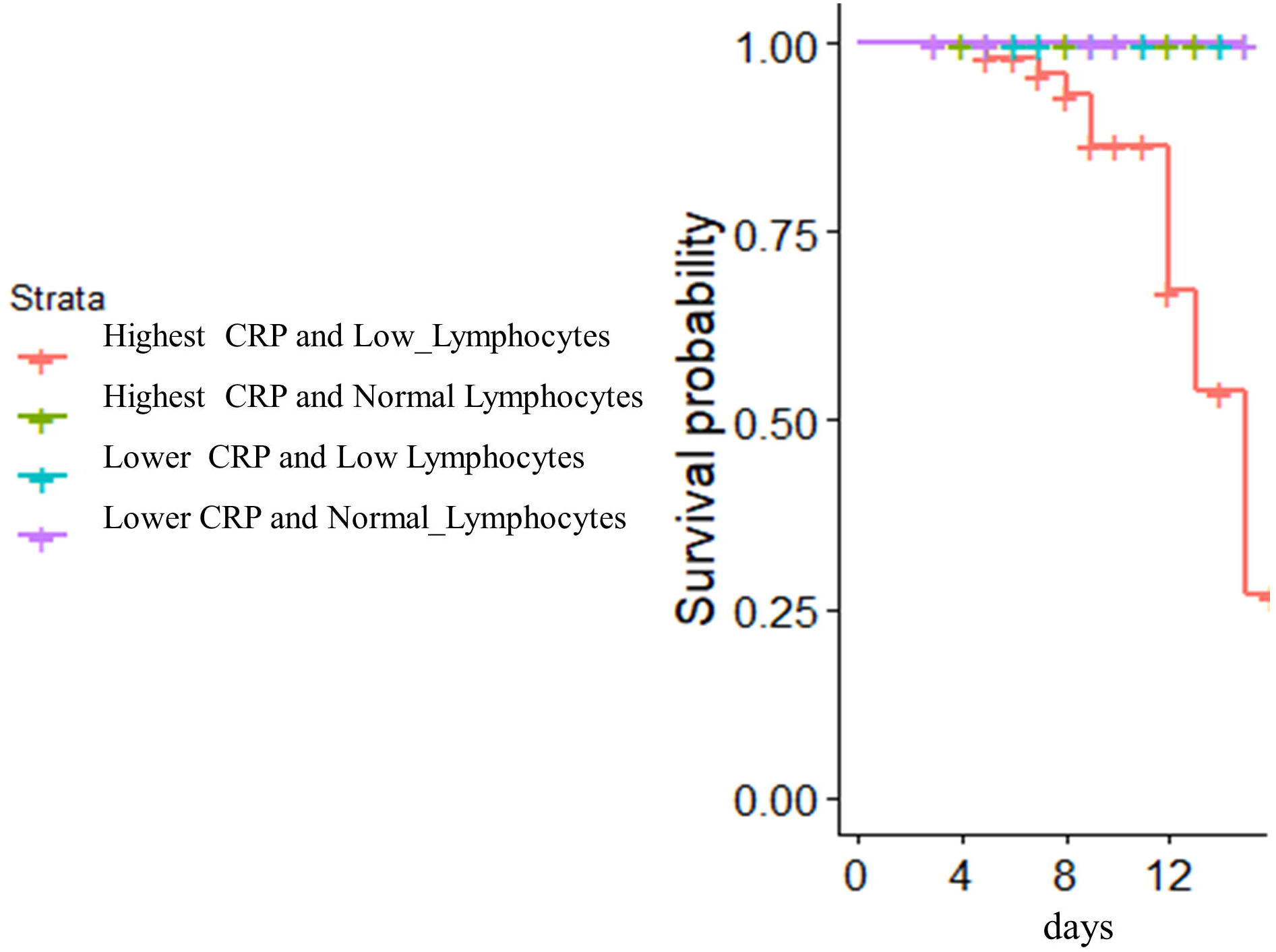
Mortality after TCZ is seen only in persons with highest CRP levels and low lymphocyte counts at treatment initiation. Highest and Lower CRP levels were > 30 and < 30 mg/L respectively. Normal and Low lymphocyte counts were > 1000 and < 1000 cells/uL respectively. There were 41 patients in the highest CRP, low lymphocyte count group (red), there were 22 patients in the highest CRP normal lymphocyte count group (green), 5 patients in the lower CRP low lymphocyte group (blue) and 6 patients in the lower CRP normal lymphocyte count group (purple).

## DISCUSSION

Here, we report the outcomes of COVID-19 disease in individuals treated with the IL-6 receptor inhibitor, Tocilizumab (TCZ).

The pathogenesis of severe COVID-19 disease is incompletely understood, however emerging evidence that includes elevated plasma levels of IL-6 and elevated levels of soluble markers that are induced by IL-6 (such as C reactive protein, ferritin and D-dimer products of fibrinolysis^10,11^ suggest that IL-6 may play an important role in the morbid outcome. In another setting of cytokine storm, i.e. the storm syndrome seen after CAR-T cell therapy IL-6 receptor blockade has been shown to be highly effective^6^

In two small series of COVID-19 cases that included 15 and 21 patients who received open label TCZ, there was an improvement in clinical course as well as in laboratory and radiographic abnormalities in some. Luo et al.^12^treated 15 patients with TCZ by vein at varying doses from 80 to 600 mg, combining TCZ with methylprednisolone in eight and repeating TCZ dosing in 5. They reported clinical improvement in one, stabilization in nine, deterioration in five patients that included three deaths. CRP levels fell in all and IL-6 levels rose in all but one patient. Increase in serum IL-6 is expected with TCZ administration as a result of receptor blockade^12^. Xu et all^13^ reported on 21 patients with severe COVID-19 pneumonia who were treated with TCZ 400 mg by vein in addition to treatment with lopinavir and methylprednisolone Clinical improvement, as characterized by rapid decrease in temperature, improvements in oxygenation and in chest CT abnormalities were reported; CRP levels fell in all patients and 19 of 21 were discharged from the hospital.

In the present study we investigated the results of TCZ treatment in a larger cohort of 89 patients with COVID-19 pneumonia. Here, we report the early effects of TCZ administration on clinical and laboratory outcomes. All our patients had SARS-2 infection as revealed by viral sequences in respiratory secretions and all had pneumonia as evidenced by CT scan results. Severity of disease varied with 17 patients requiring mechanical ventilation support before administration of TCZ. Although all patients were treated with drugs often used for treatment of SARS-2 infection such as antimalarials (chloroquine or hydroxychloroquine), azithromycin or an antiviral protease inhibitor (lopinavir/ritonavir), none of these agents has confirmed clinical utility in treatment of COVID-19^14,15^. In contrast to the two smaller series cited above^16,13^ corticosteroids were not given to these patients during TCZ administration or in the 2 days following. The role of corticosteroids in treatment of COVID-19 is unproven^17,18^.

Our patients with COVID-19 pneumonia had a profound inflammatory disease. Specifically, all patients had elevated levels of C reactive protein exceeding the 6 mg/L upper limit of normal in our laboratories. Plasma IL-6 levels also were elevated in all patients exceeding the laboratory normal reference range of 6.4 pg/mL and circulating lymphocyte numbers were diminished in most patients. The drivers of inflammation and lymphocytopenia in COVID-19 are not well understood yet but the preliminary data generated to date by this and other^16, 13^ studies suggest that interfering with IL-6 activity improves these indices. The rapid improvement in NEWS2 scores after TCZ administration suggests that clinical improvement is linked to resolution of inflammation and that IL-6 activity is an important driver of morbidity in SARS-2 infection. In our series, outcome of disease in treated patients who did not require mechanical ventilation was largely favorable with nearly 90% of patients surviving to discharge and only one death among 72 treated patients at the time of writing. Clinical outcomes in patients who were receiving mechanical ventilation at initiation of TCZ treatment were not as good. Although 10 of 17 patients receiving mechanical ventilation experienced an improved NEWS2 score 1–2 days after TCZ treatment. (4 of these 10 patients died), overall there were 10 deaths in the entire group, while 7 patients remained in hospital.

While lymphocytopenia and elevated C reactive protein are common in COVID-19, mortality was only seen in patients who had both low lymphocyte counts (< 1000/uL) and profoundly elevated CRP (>30 mg/L) before treatment with TCZ. In this regard, a recent case series of 209 persons with COVID-19 found that elevated levels of C reactive protein, particularly levels exceeding 26.9 mg/L predicted clinical deterioration. Lymphocytopenia was not associated with morbid deterioration but those who progressed had lower lymphocyte proportions than those who did not progress^19^.

Although our results suggest that TCZ therapy is beneficial for hospitalized patients with COVID-19, this report has important limitations. First, this was a retrospective study conducted in a heterogeneous group of patients. Interpretation of this study is also limited by the absence of a control group that did not receive TCZ. Nonetheless the temporal improvement in clinical and laboratory scores suggest that these are related to TCZ administration. Controlled studies are needed to determine whether cytokine blockade and in particular, blockade of IL-6 confers benefit to persons with COVID-19. Such studies are ongoing and their findings are eagerly awaited. Our data suggest that for COVID-19 patients requiring mechanical ventilation, IL-6 blockade alone may be insufficient; other interventions are necessary.

## Data Availability

The data referred to in this ms are contained in the hospital records of City Clinical Hospital No.52 of Moscow Healthcare Department, Moscow, Russia

## Declaration of interest

We declare no competing interest

## Acknowledgements

We would like to thank Dr. Leonid Margolis for encouragement and for assistance in manuscript preparation. We would thank Dr. Alexey Mazus for encouragement and help in coordinating this research

## Notes

### Competing Interest Statement

The authors have declared no competing interest.

### Funding Statement

No external funding

### Author Declarations

The study was approved by the local ethics committee of Moscow City Clinical Hospital No. 52

